# Macular vascular complexity analysis of diabetes mellitus by swept-source optical coherence tomographic angiography

**DOI:** 10.1101/2021.08.16.21262124

**Authors:** Jiahui Liu, Shuhui Chen, Zhiyi Xu, Wei Wang, Dingqiao Wang, Yongyue Su, Shulun Zhao, Meichan Li, Fengling Chen, Chengguo Zuo, Minyu Chen

## Abstract

**Purpose:** To evaluate the associations of retinal vascular complexity features, including fractal dimension (FD) and blood vessel tortuosity (BVT), with the severity of diabetic retinopathy (DR) by using noninvasive optical coherence tomographic angiography (OCTA).

**Methods:** This prospective cross-sectional study recruited ocular-treatment-naïve patients with type 2 diabetes mellitus (DM) registered in the community of Guangzhou, China. OCTA was used to obtain the measurements of FD and BVT in the superficial capillary plexus (SCP) and the deep capillary plexus (DCP). Univariate and multivariate linear regression was used to analyse the correlation of FD and BVT in different layers with DR severity.

**Results:** A total of 1282 patients with DM (1282 eyes), with a mean age of 64.2±7.8 years, were included. FD in the DCP decreased and BVT in the DCP increased in DR patients compared to non-DR patients, even after adjusting for confounding factors (P< 0.05). Trend analysis showed a significant decrease in the FD values as the DR progressed, while the BVT progressively increased with worsening DR severity (P< 0.01). The FD in DCP had a statistically significant positive correlation with FD in SCP and a negative correlation with BVT in SCP and BVT in DCP in all of the participants, including the non-DR group, moderate DR group and severe DR group (P< 0.01).

**Conclusion:** FD and BVT determined using OCTA might be useful parameters for objectively distinguishing DR from non-DR and indicating DR progression.

## Introduction

Diabetic retinopathy (DR) is the leading cause of blindness in the working-age population around the world ^1-3^. Patients with diabetes mellitus (DM) are usually symptomless until the advanced stages of DR, leading to a poor visual prognosis after treatment. Thus, early detection and prompt treatment of DR play a pivotal role in protecting DM patients from vision impairment. Although the exact pathogenesis of DR remains unclear, it is universally acknowledged that damage to the retinal microvasculature, especially in the macula, might contribute to the occurrence and development of DR^4, 5^. Capillary drop-out has been reported to precede proliferative disease ^6^. However, there are no available clinically quantitative parameters to identify the degree of microvascular damage, especially in the early stage of DR.

As optical coherence tomography angiography (OCTA) has become more widely available, detailed microvascular networks can now be quantified conveniently and noninvasively with high resolution. With OCTA, some abnormal changes in the retinal capillary network in DM patients without DR have been recognized, demonstrating that OCTA metrics could be early indicators of subtle, subclinical alterations in DM before the first clinically observable signs of DR ^7-9^. Fractal dimension (FD) and blood vessel tortuosity (BVT) have been used to evaluate the overall structural and geometric complexity of retinal vasculatures. Recent studies revealed that FD and BVT were altered in DR and therefore were DR biomarker candidates ^2, 3, 10, 11^. However, these findings did not consider confounding factors, such as age, sex, blood pressure, HbA1c or OCTA signal strength intensity. In addition, the correlation of FD and BVT has not been elucidated to date. We are unaware of any studies about vascular complexity features in diabetes patients from Chinese communities. Therefore, the objective of this study was to assess FD and BVT to identify localized complexity distortions across various stages of DR with OCTA and to determine the relationship between them in a large sample of Chinese patients with DM.

## Methods

### Participants

This prospective cross-sectional study was conducted in Zhongshan Ophthalmic Center (ZOC) with the permission of the Ethics Committee of the ZOC (2017KYPJ094). All investigations followed the tenets of the Declaration of Helsinki. Subjects in the study were enrolled from November 01, 2017, to December 30, 2020, in the community of Guangzhou, China. Patients aged 35–80 years with type 2 diabetes and without any history of eye treatments or surgeries (ocular treatment naïve) were included in this study. The exclusion criteria were as follows: (1) eyes with other significant ocular pathology (including hypertensive retinopathy, retinal vascular occlusions, and uveitis); (2) history of other systemic diseases, such as malignant tumours, ischaemic heart disease, stroke, cancer, or kidney disease; (3) a history of heart bypass, thrombolysis, or kidney transplantation; cognitive impairments or mental illnesses that made them unable to complete questionnaires or cooperate with the examiner; (4) spherical equivalent (SE) > −12.0 D, astigmatism > +4.0 D, axial length (AL) > 26.0 mm; (5) a best corrected visual acuity (BCVA) less than 20/200; (6) abnormal refractive media (such as moderate to severe cataract, corneal ulcer, or severe pterygium), poor fixation or other causes that lead to poor quality of fundus, OCT or OCTA images. When both eyes were eligible for the study, the worse eye graded by DR from each patient was selected as the study eye. When both eyes had an equal grade of DR, the right eye was selected as the study eye.

### General and laboratory parameters

Demographic and systemic information, including age, sex, and medical history, was obtained using standardized questionnaires, and the information was confirmed after further inspection of outpatient history. Height, weight, systolic blood pressure (SBP), and diastolic blood pressure (DBP) were measured using standardized processes. Haemoglobin A1c (HbA1c), total cholesterol and triglycerides were measured in a laboratory certified by the Chinese government.

### Ocular examination

Each participant underwent a full clinical examination, including slit-lamp bioscopy, ophthalmoscopy, intraocular pressure (IOP), visual acuity, refraction, ocular biometry, retinal photography, and OCTA imaging. Ocular biometric parameters were measured with a Lenstar LS900 (Haag-Streit AG, Koeniz, Switzerland), including AL, corneal curvature, lens thickness and anterior chamber depth. Pupil dilation was performed with instillation of 0.5% tropicamide plus 0.5% phenylephrine in eye drops. Once the pupils were fully dilated, standardized 7-field colour retinal photographs adhering to the Early Treatment of Diabetic Retinopathy Study (ETDRS) protocol were taken using a digital fundus camera (Canon CR-2, Tokyo, Japan). The clinician retinopathy severity grading was used for patient recruitment. For the analysis, a masked grading of the colour photographs assigned eyes into the non-DR group (no retinopathy), mild DR group (mild nonproliferative diabetic retinopathy), moderate DR group (moderate nonproliferative diabetic retinopathy), and severe DR group (including severe nonproliferative diabetic retinopathy and proliferative DR) for final analysis by two trained ophthalmologists and if there was any disagreement about the grade, it was confirmed by a senior retina specialist ^12^.

### OCTA imaging and measurements

A commercial swept-source OCTA instrument (DRI OCT-2 Triton; Topcon, Tokyo, Japan) was used for retinal microvasculature imaging. It had a central wavelength of 1050 nm with an axial resolution of 8 μm, a transverse resolution of 20 μm, and a speed of 100,000 A-scans per second. We used 3 mm × 3 mm scans that provided a more detailed description of the fovea and its contour. Automated segmentation and image exportation generated SS-OCTA images of the superficial capillary plexus (SCP) and the deep capillary plexus (DCP). Once moderate segmentation errors appeared, we corrected the segmentation manually. All OCTA images were acquired by a well-trained examiner with no knowledge of the study protocol. The image quality of each scan was automatically graded by the built-in software, with a signal strength index (SSI) ranging from 0 to 100 ^13^. Images with SSI≤50 were included.

We measured two complexity quantitative features, FD and BVT, in the OCTA images. The OCTA images were standardized, cropped, and binarized using Fiji (free downloadable software, https://fiji.sc) ^14^. Before measuring the OCTA features, a binary vessel map and skeleton map were extracted from the input images. FD can quantify the degree of branching complexity (i.e., blood vessels). In our study, the box counting technique was used to measure the FD values in both SCP and DCP^15^. The FD analysis procedure has been described in a previous study ^15^. The average BVT was measured as the ratio of the sum of the actual branch lengths to the sum of the straight lengths between the branch nodes^16^. The FD and BVT were measured using segmented branches within the skeleton map in the SCP and DCP (Figure 1). As the FD and BVT values were too small, we enlarged their values by 1000 times for statistical analysis.

**Figure 1.**
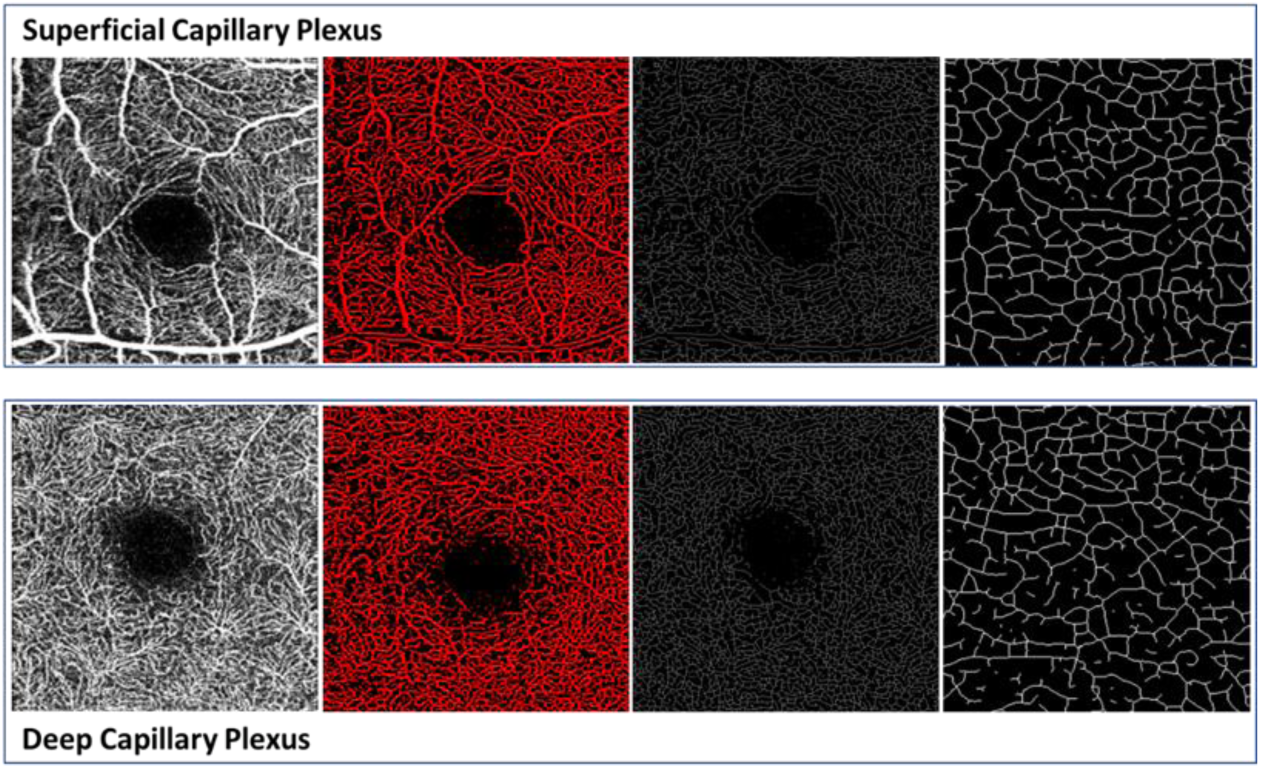
Illustration of fractal dimension (FD) and blood vessel tortuosity (BVT) measurements in the superficial capillary plexus (SCP) and deep capillary plexus (DCP) by OCTA. Column 1: OCTA image. Column 2: Segmented blood vessel map including large blood vessels and small capillaries. Column 3: Skeletonized blood vessel map. Column 4: Image from Column 3 magnified by a thousand times.

### Statistical analysis

The Shapiro–Wilk test was used to verify the normal distribution, and analysis of variance (ANOVA) was performed to compare the differences in the demographic, systemic, and ocular parameters among the groups with different stages of DR. Categorical variables were analysed by χ2. Linear regression analysis was used to evaluate the association between FD in DCP and the other OCTA features among the different groups. Univariate and multivariate linear regressions were used to compare FD and BVT in both SCP and DCP between the control group and the various DR groups. Model 1 of the multivariable linear regression was adjusted for age, sex and OCTA signal strength intensity. Model 2 was performed by adjusting for more confounding factors, including age, sex, systolic blood pressure, diastolic blood pressure, HbA1c, serum creatinine, blood cholesterol, triglycerides, axial length, and OCTA signal strength intensity. All analyses were performed using Stata Version 16.0 (Stata Corporation, College Station, TX, USA). A P value < 0.05 was considered statistically significant.

## Results

### Demographics

A total of 1282 eyes from 1282 subjects were included in the study. The demographic and clinical characteristics of the study participants are shown in Table 1. The average age was 64.2±7.8 years, and 756 (58.97%) patients were male. Of the 1059 patients without DR, the mean age was 64.3 ± 7.5 years, and 644 (60.81%) were male. Of the 54 patients with mild DR, the mean age was 62.8 ± 6.9 years, and 30 (55.56%) were male. Of the 133 patients with moderate DR, the mean age was 63.8 ± 9.3 years, and 64 (48.12%) were male. Of the 36 patients with severe DR, the mean age was 62.1 ± 8.8 years, and 18 (50%) were male. The parameters relating to sex, SBP, HbA1c, serum creatinine and OCTA macular signal strength intensity differed among the groups (P < 0.05). No statistically significant differences were present with respect to the other parameters.

**Table 1.** Demographics and clinical characteristics of the study participants.

| Characteristics | ALL | Non-DR | Mild DR | Moderate DR | Severe DR | p Value |
| --- | --- | --- | --- | --- | --- | --- |
| No. of subjects | 1282 | 1059 | 54 | 133 | 36 |  |
| Male (%) | 756(58.97%) | 644(60.81%) | 30(55.56%) | 64(48.12%) | 18(50%) | 0.024 |
| Mean age, year | 64.2±7.8 | 64.3±7.5 | 62.8±6.9 | 63.8±9.3 | 62.1±8.8 | 0.182 |
| Systolic blood pressure, mm Hg | 135.5±19.0 | 134.9±18.8 | 133.3±16.0 | 138.8±19.3 | 145.3±24.2 | 0.002 |
| Diastolic blood pressure, mm Hg | 70.9±10.3 | 71.1±10.1 | 70.9±12.0 | 69.0±10.1 | 70.8±12.7 | 0.161 |
| HbA1c, % | 7.0±1.5 | 6.8±1.3 | 7.9±1.9 | 7.9±1.7 | 8.6±1.8 | < 0.001 |
| Triglycerides, mmol/L | 2.3±1.6 | 2.3±1.6 | 2.2±1.6 | 2.3±1.9 | 2.2±1.3 | 0.919 |
| Total cholesterol, mmol/L | 4.8±1.0 | 4.9±1.0 | 4.8±1.1 | 4.8±1.1 | 4.9±1.1 | 0.691 |
| Serum creatinine, µmol/L | 70.9±21.1 | 69.3±19.1 | 72.6±23.7 | 79.4±28.9 | 82.3±28.0 | < 0.001 |
| Axial length, mm | 23.4±0.9 | 23.4±0.9 | 23.2±0.9 | 23.2±0.8 | 23.2±0.9 | 0.136 |
| OCTA signal strength intensity | 59.2±11.5 | 60.0±11.1 | 59.5±12.3 | 55.7±12.3 | 49.5±14.1 | < 0.001 |

### Distribution of FD and BVT by DR severity

A summary of the average and detailed univariate analysis of FD and BVT in SCP and DCP between the control group and the different DR groups is shown in Table 2 and Figure 2.

**Table 2.**
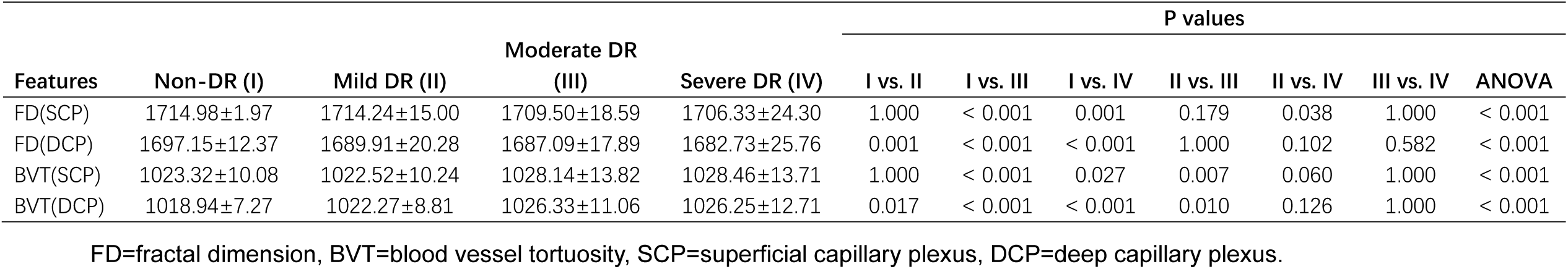
Comparisons of FD and BVT in different stages of diabetic retinopathy.

**Figure 2.**
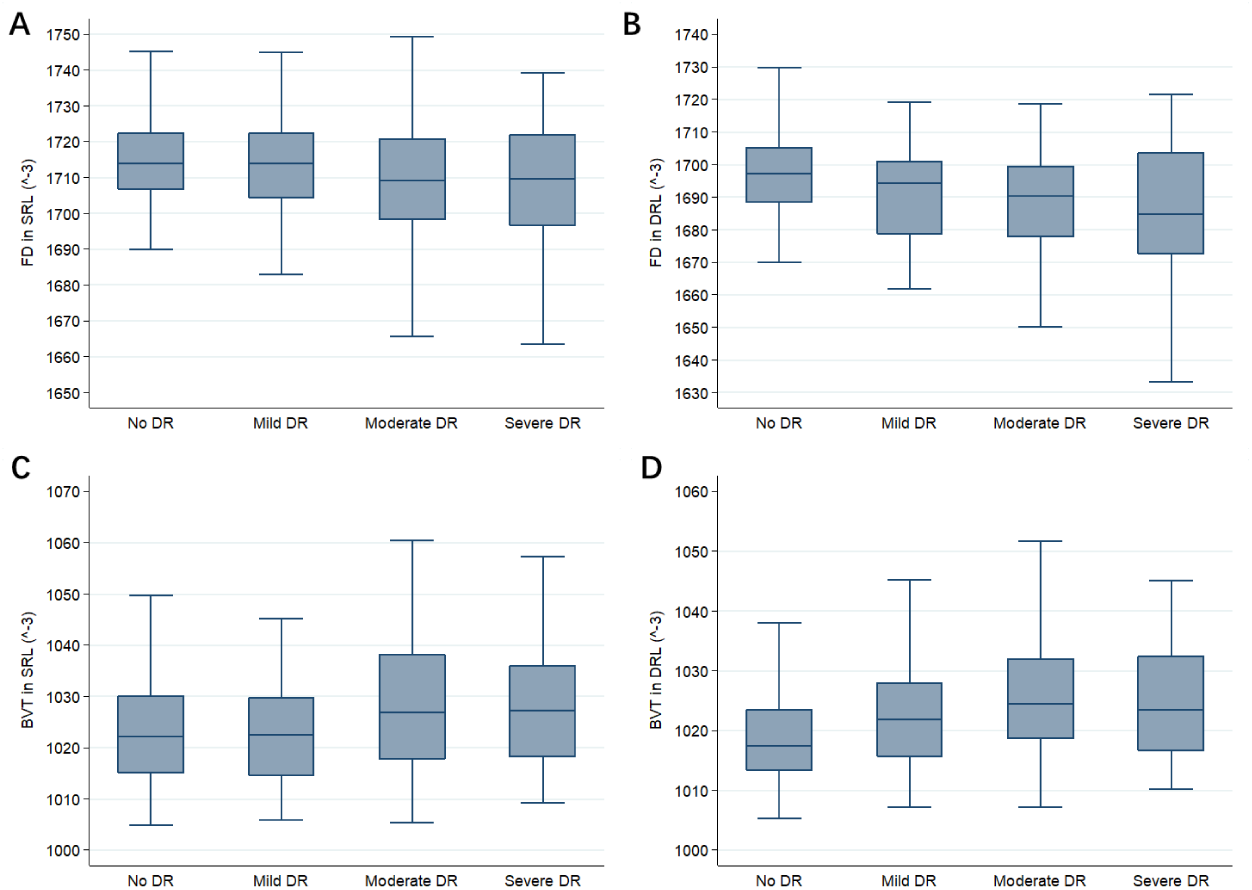
Boxplots showing the distribution of fractal dimension (FD) and blood vessel tortuosity (BVT) in the superficial retinal layer (SRL) and the deep retinal layer (DRL) by swept-source optical coherence tomographic angiography stratified by various stages of DR. (A) Average FD in the SRL of various stages of DR; (B) Average FD in the DRL of various stages of DR; (C) Average BVT in the SRL of various stages of DR; (B) Average BVT in the DRL of various stages of DR.

A post hoc study after ANOVA showed that all of the features were able to distinguish controls versus moderate DR patients (P< 0.001) and controls versus severe DR patients (P< 0.05), but none of the features showed statistically significant differences between moderate DR eyes and severe DR eyes. Comparison of FD and BVT in DCP between non-DR patients and mild DR patients showed a significant difference (P< 0.05). Furthermore, there was a significant increase in BVT in SCP and in DCP of mild DR eyes compared with moderate DR eyes (P ≤ 0.01). Only FD in SCP was able to distinguish between the mild DR group and the severe DR group (P=0.038). The FD in DCP was higher in non-DR eyes than in DR eyes at each stage (P ≤ 0.001), but there was no significant difference among the different stages of DR. Consequently, we considered that FD in DCP was an important feature to differentiate DR and non-DR.

### Association between Vascular Complexity and Diabetic Retinopathy

A univariate linear regression analysis was performed to observe the relationship between FD in DCP and other OCTA features (Table 3). FD in DCP had a statistically significant positive correlation with FD in SCP and a negative correlation with BVT in SCP and BVT in DCP in all of the participants, non-DR group, moderate DR group and severe DR group (P< 0.01). Only BVT in DCP showed a negative trend with FD in DCP in mild DR eyes (P< 0.001).

**Table 3.**
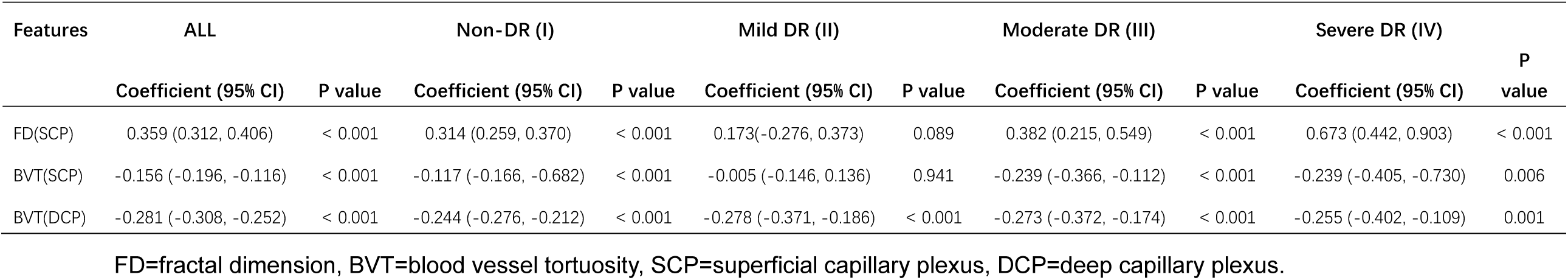
Univariate linear regression analysis of the relationship between features of vascular complexity and FD in DCP.

**Table 4.** Multivariable linear regression of OCTA parameters in non-DR eyes compared with various stages of DR eyes after adjusting for confounding factors.

| Features | Non-DR vs. Mild DR<br>Coefficient (95% CI) | P value | Non-DR vs. Moderate DR<br>Coefficient (95% CI) | P value | Non-DR vs. Severe DR<br>Coefficient (95% CI) | P value |
| --- | --- | --- | --- | --- | --- | --- |
| Model 1 |  |  |  |  |  |  |
| FD(SCP) | -0.43(-3.59, 3.09) | 0.810 | -6.72(-9.05, -4.40) | < 0.001 | -13.63(-17.98, -9.27) | < 0.001 |
| FD(DCP) | -7.96(-11.91, -4.01) | < 0.001 | -10.19(-12.79, -7.59) | < 0.001 | -16.26(-21.15, -11.37) | < 0.001 |
| BVT(SCP) | -1.01(-4.04, 2.03) | 0.516 | 4.65(2.65, 6.64) | < 0.001 | 6.48(2.72, 10.24) | 0.001 |
| BVT(DCP) | 3.58(1.29, 5.86) | 0.002 | 7.06(5.55, 8.57) | < 0.001 | 7.90(5.08, 10.73) | < 0.001 |
| Model 2 |  |  |  |  |  |  |
| FD(SCP) | -0.89(-4.49, 2.72) | 0.630 | -6.72(-9.15, -4.28) | < 0.001 | -13.74(-18.31, -9.17) | < 0.001 |
| FD(DCP) | -6.75(-10.72, -2.78) | 0.001 | -8.93(-11.62, -6.25) | < 0.001 | -13.37(-18.40, -8.34) | < 0.001 |
| BVT(SCP) | -0.61(-3.71, 2.49) | 0.700 | 4.62(2.53, 6.72) | < 0.001 | 6.63(2.70, 10.55) | 0.001 |
| BVT(DCP) | 3.55(1.23, 5.87) | 0.003 | 6.66(5.09, 8.23) | < 0.001 | 7.43(4.49, 10.37) | < 0.001 |

Model 1 was adjusted for age, sex and OCTA signal strength intensity. Model 2 was further adjusted for confounding factors, including age, sex, systolic blood pressure, diastolic blood pressure, HbA1c, serum creatinine, blood cholesterol, triglycerides, axial length, and OCTA signal strength intensity. FD=fractal dimension, BVT=blood vessel tortuosity, SCP=superficial capillary plexus, DCP=deep capillary plexus.

To reduce the factors known to be relevant to OCTA features, we performed two multivariate linear regression analyses to compare FD and BVT in both SCP and DCP between the control group and the various DR groups, which was adjusted for confounding factors. The results of the two models were consistent with the univariate analysis in Table 2, but nearly all of the differences between groups were of a greater magnitude.

## Discussion

This OCTA study based on a large sample demonstrated that FD in DCP decreased and BVT in DCP increased in various grades of DR compared to non-DR. The differences were of a greater magnitude after adjusting for potential confounding factors. In addition, our results showed that FD in DCP was positively correlated with FD in SCP and negatively correlated with BVT in SCP and BVT in DCP in all groups except the mild DR group. These findings confirmed a close relationship between alterations in the retinal microcirculation, particularly in DCP, and the pathophysiology of DR. To the best of our knowledge, this study is the first to use SS-OCTA to investigate the association of vascular complexity features, including FD and BVT, and different stages of DR in patients with diabetes from communities in China.

Fractal analysis has been employed as a useful tool to study the retinal vascular pattern and measure the complexity of retinal vasculature branching. As DR develops, the high rate of vascular dropouts results in a lower vessel density, which is represented by a lower average number of FD values^3^ [9]. Chen et al. reported that the value of FD in the four quadrants of the deep retinal capillary layer was lower in diabetic patients with minimal DR than in those with no DR, and FD in the deep retinal capillary layer was much lower in DM eyes without DR than in nondiabetic control subjects^17^. Suruchi et al. showed that FD in both the superficial and deep capillary plexuses of diabetic eyes was significantly lower than that in control eyes^10^. Consistent with previous studies, our findings illustrated that retinal vascular FD in both SCP and DCP showed a significant reduction in DR patients. Moreover, the superiority of our research was that we adjusted for a large number of confounding factors associated with vascular complexity. However, no statistically significant changes were observed in FD in either the superficial or deep vascular networks among the different stages of DR, apart from FD at SCP in the severe DR group compared with the moderate DR group. These results suggested that FD in DCP could be an important indicator that distinguishes DR from non-DR, indicating that the early changes of retinal capillaries associated with DR may first occur in the deep retinal capillary layer before extending to the superficial plexus.

There was no significant difference in FD of the superficial capillary plexus when comparing mild DR and non-DR, suggesting that the superficial capillary plexus may be damaged until DR progresses to a greater degree. This is further supported by our analysis of BVT, which did not yield a significant difference in the superficial capillary plexus between control eyes and mild DR eyes. Similar to the FD in DCP, the BVT in DCP in non-DR was lower than in any other stages of DR, even after adjusting for all known covariates. This result is consistent with a previous study by Minhaj et al. ^3^. Consequently, BVT in DCP could be used for non-DR vs. DR classification.

Retinal vessel tortuosity has been considered a characteristic of haemodynamic changes and grading of DR ^18-20^. Dysfunction of the endothelium and pericytes induces unstable vessel walls, which can easily become tortuous^21, 22^. Neovascularization often occurs in DR patients and leads to an increase in BVT^20^. However, neither FD nor BVT in DCP distinguishes between different severities of DR, which is inconsistent with Suruchi’s^10^ and Hyungwoo’s research^16^. This may be due to the small sample size of the enrolled DR populations or heterogeneous study populations.

As FD in DCP plays a vital role in the diagnosis and risk assessment of DR, we performed univariate linear regression analyses between FD in DCP and other OCTA features. Few studies have analysed the association between FD and BVT. It was shown in our correlation analysis that the negative associations of FD in DCP with both BVT in SCP and DCP are plausible because all of these parameters are related to the abnormal vascular architecture with DR progression. As DR progresses, avascular regions are assigned small FD values, and interrupted blood flow results in short branches. Short capillaries are prone to become tortuous rather than long branches. The cerebral microvascular meshwork demonstrated that vessels closer to the capillary network have higher tortuosity and more curvature^23^, which supports our deduction. Additional studies are necessary to estimate the cause of high tortuosity by short branches.

An advantage of our study is that all participants were treatment-naïve patients with type 2 DM from a single Chinese community, consisting of a homogenous and large-scale cohort. In addition, the model was adjusted for several potential confounding factors, making our results more accurate and reliable. There are also some limitations to this study. First, our observations were only of a very small area of the retina because the 3 × 3 mm area showed more clearly demarcated vessels, ensuring more precise binarization of vessels and, subsequently, more accurate quantification of the parameters. A larger scan area could be performed to quantify OCTA features in more regions of the retina. Second, projection and motion artefacts in OCTA and blood flow velocity below the threshold value can affect quantitative feature analysis. We strictly controlled the image quality by a quality index. Third, we did not have enough patients in each stage of DR because the participants in our study were recruited from communities. Further work is warranted with a larger sample size of DR patients. Finally, this study, as a cross-sectional study, cannot illustrate the cause-effect relationship between vascular alterations and DR occurrence and development. Longitudinal studies are required in the future.

## Conclusions

In summary, we found that FD and BVT in DCP by OCTA imaging may be new quantitative biomarkers for the diagnosis and risk assessment of DR. Future studies are necessary to determine whether intensive investigations of retinal vascular geometry can benefit patients and improve clinical decision-making in the risk evaluation and management of DR.

## Data Availability

Data can be available when approprieate request.

## Disclosures

Financial Disclosures: None.

## Notes

**Funding information:** This research was supported by the National Natural Science Foundation of China (82000901, 82102993), the Fundamental Research Funds of the State Key Laboratory of Ophthalmology (303060202400362), Guangdong Basic and Applied Basic Research Foundation (2019A1515110618) and the Research and Development Foundation of Dongguan People’s Hospital (201900181, K202008).

### Competing Interest Statement

The authors have declared no competing interest.

### Clinical Trial

ISRCTN registry no: 15853192

### Funding Statement

This research was supported by the National Natural Science Foundation of China (82000901), the Fundamental Research Funds of the State Key Laboratory of Ophthalmology (303060202400362).

### Author Declarations

Medical Ethics Committee of the Zhongshan Ophthalmic Centre approved the study (2017KYPJ094)

